# Impact of HIV Self-Testing on Recent HIV Testing Among Women in Uganda: A Propensity Score Matched Analysis Using the 2022 UDHS

**DOI:** 10.64898/2026.08.04.26359666

**Authors:** Geofrey Emesu, Solome Najjuma, Persis Tikabulamu, Aggrey David Mukoose, Joseph Kagaayi

## Abstract

**Background:** HIV self-testing (HIVST) has been promoted as a strategy to reach individuals who do not access facility-based testing. However, evidence on whether HIVST leads to more frequent testing among women of reproductive age in Uganda remains limited. This study evaluated the impact of HIV self-testing on recent HIV testing among women using nationally representative data.

**Methods:** Data were drawn from the 2022 Uganda Demographic and Health Survey (UDHS), including 6,438 women aged 15–49 years. The primary outcome was recent HIV testing, defined as having tested for HIV within the 12 months preceding the survey. The treatment variable was ever having used HIV self-testing. Propensity score matching (PSM) with 1:1 nearest neighbour matching (caliper = 0.05) was used to balance observed covariates including parity, media exposure, education, residence, wealth quintile, health insurance, and age group. The average treatment effect on the treated (ATT) was estimated.

**Results:** Among 6,438 women, 23.87% (1,537) reported ever using HIV self-testing. Recent HIV testing was observed in 67.4% of HIVST users compared to 47.4% of non-users (unmatched difference = 20%). After matching, HIVST use increased the likelihood of recent testing by 15.6 percent (ATT = 15.6%; SE = 0.052; t = 2.99). Covariate balance was achieved post-matching, with mean bias reduced from 20.5% to 0.7%, and the B statistic falling from 50.4% to 2.5% (below the 25% threshold). All standardized differences were substantially reduced, with education showing perfect balance (100% reduction) and wealth showing 97.6% reduction.

**Conclusion:** HIV self-testing significantly increases recent HIV testing among women of reproductive age in Uganda. Expanding access to HIVST, particularly for women with lower education, those in poorer wealth quintiles, and those without media exposure, could improve testing frequency and support progress toward the UNAIDS 95-95-95 targets.

## Introduction

The global HIV epidemic remains a major public health challenge in sub-Saharan Africa, with Uganda recording an adult HIV prevalence of 5.2% as of 2022 (1). Women of reproductive age bear a disproportionate burden of HIV infection. In Uganda, HIV prevalence among women aged 15–49 years is approximately 6.5%, compared to 4.0% among men (2). This disparity reflects a combination of biological vulnerability, gender inequalities, socioeconomic disadvantage, and limited access to sexual and reproductive health services (3). Regular HIV testing is critical for reducing transmission, preventing mother-to-child transmission, initiating antiretroviral therapy promptly, and achieving epidemic control. The UNAIDS 95-95-95 targets require that 95% of people living with HIV know their status. Yet, a substantial proportion of women in Uganda either never test or test infrequently.

Conventional facility-based HIV testing has been the mainstay of diagnosis for decades. However, barriers for women including stigma, transport costs, long waiting times, fear of breach of confidentiality, lack of child care, and concerns about partner notification. These challenges disproportionately affect women who experience significantly higher vulnerability due to gender inequalities, socioeconomic disadvantage, and power imbalances within relationships (3).

HIV self-testing (HIVST) emerged as an alternative approach, allowing individuals to test privately and receive results within 20 minutes (4). Since WHO recommended HIVST in 2016 (5), Uganda has piloted and scaled up distribution through antenatal care, community-based programmers, and private pharmacies (6). For women, HIVST offers particular advantages: it can be used discreetly, shared with partners, and integrated into existing maternal health contacts (7). Recent studies in sub-Saharan Africa have demonstrated increasing uptake and acceptability of HIV self-testing among women due to its convenience, confidentiality, and flexibility (8) and evidence from cross-sectional studies in urban Uganda has shown that HIV self-testing increases recent and frequent HIV testing among female sex workers (9), suggesting that self-testing can overcome barriers to facility-based testing such as stigma, facility hours, and transportation costs.

The key policy question is whether HIVST actually achieves its intended goal among women by more frequent and regular testing is the pathway to early diagnosis and prevention of mother-to-child transmission. The UDHS measures time since last HIV test, which is a strong proxy for testing behavior. Women who test every 12 months or less are more likely to be diagnosed early if they seroconvert and are more likely to receive prevention of mother-to-child transmission services if they become pregnant.

Some studies have also shown associations between HIVST and testing frequency(10, 11), but few have focused specifically on women or used rigorous causal methods. Women who self-test may differ systematically from those who test at facilities in terms of education, parity, antenatal care attendance, or prior testing experience. Without adjusting for these confounders, estimates of impact are likely biased (12). A systematic review and meta-analysis found that HIVST increased testing uptake and improved linkage to care (5), but evidence specific to women in Uganda remains limited. A recent multi-country analysis of 21 sub-Saharan African countries found that knowledge and utilization of HIVST among women was only 2.17%, with education, wealth, media exposure, and prior HIV testing history identified as significant predictors(13).

In this study, we applied propensity score matching (PSM) to the 2022 Uganda Demographic and Health Survey (UDHS) to estimate the causal effect of HIV self-testing on recent HIV testing (within 12 months) among women of reproductive age. PSM allows us to balance observed covariates between HIVST users and facility-based testers, mimicking randomization and providing a credible estimate of the average treatment effect on the treated (14)

## Materials and Methods

### Study Design and Data Source

This was a secondary analysis of cross-sectional data from the 2022 Uganda Demographic and Health Survey (UDHS). The UDHS is a nationally representative household survey conducted every five years by the Uganda Bureau of Statistics (UBOS) in collaboration with the Ministry of Health and ICF International (UBOS & ICF, 2023). The survey used a two-stage stratified cluster sampling design. In the first stage, enumeration areas (EAs) were selected with probability proportional to size. In the second stage, households were systematically sampled from each EA. All women aged 15–49 years in selected households were eligible for individual interviews.

#### Study Population

We included women aged 15–49 years who reported ever having been tested for HIV. Women with missing data on testing method (self-test vs. facility) or outcome (time since last test) were excluded. The final analytical sample for propensity score matching comprised of 6,438 women.

#### Variables

**Outcome variable**: Recent HIV testing was defined as having tested for HIV within the 12 months preceding the survey. This variable is derived from the UDHS question: “ When was your most recent HIV test?”(q1025y) Responses ranged from 2000 to 2022. We coded recent testing as 1 if the test occurred within the last 12 months, and 0 otherwise. This threshold aligns with WHO guidance on annual testing for populations at ongoing risk, including women of reproductive age.

**Treatment variable**: HIV self-testing (HIVST) was coded as 1 if the woman reported using a self-test kit for her most recent HIV test, and 0 if the test was conducted at a health facility (including government, private, or NGO facilities). Home-based testing by a community health worker was not classified as self-testing.

**Covariates:** Matching covariates were selected based on prior literature and theoretical considerations (15) and included: age group (15–24, 25–34, 35–49 years), highest education level (none, primary, secondary, higher), wealth quintile (poorest, poorer, middle, richer, richest), type of place of residence (urban/rural), parity (0, 1–2, 3–4, 5 or more children), health insurance coverage (yes/no), and exposure to mass media (yes/no, defined as reading a newspaper, listening to radio, or watching television at least once a week).

#### Propensity Score Matching

Propensity scores were generated using a multivariate logistic regression model including covariates listed above. The propensity score represented the predicted probability of using HIV self-testing given observed characteristics(16). The choice of variables for the propensity score model was guided by prior literature and the theoretical framework of Andersen’s Behavioural Model of Health Services Use (17), which posits that health services utilization is influenced by predisposing (e.g., age, education, marital status), enabling (e.g., wealth, residence, health insurance), and need factors (e.g., prior testing history).

We then performed 1:1 nearest neighbor matching without replacement, using a caliper of 0.05 to restrict the maximum permitted difference in propensity scores between matched pairs. This caliper width is conservative and follows recommendations in the literature (12, 18).. A caliper of this magnitude ensures that matches are of high quality by preventing the matching of individuals with substantially different propensity scores (19). Women outside the region of common support were excluded (20). The matching was performed using the psmatch2 module in Stata (21).

#### Balance Assessment

Balance between treated (HIVST users) and untreated (non-users) groups was assessed before and after matching using standardized differences. A standardized difference of less than 0.10 indicated good balance (20, 22).. This measure is preferred over hypothesis testing because it is independent of sample size and provides a direct measure of the magnitude of differences between groups (23). We also compared variance ratios (target 0.5–2.0), visually inspected density plots of propensity scores, and assessed the overall balance using the B statistic (standardized difference of the means of the propensity scores, target < 25%) and the R statistic (ratio of the variances of the propensity scores, target 0.5–2.0)

#### Treatment Effect Estimation

The average treatment effect on the treated (ATT) was estimated as:

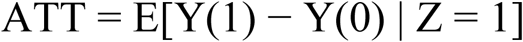

where Y(1) is the outcome (recent HIV testing) for an HIVST user, Y(0) is the counterfactual outcome had that same woman used facility-based testing, and Z indicates treatment status. Standard errors were obtained through bootstrapping with 150 replications to account for the fact that the propensity score is estimated. The number needed to treat (NNT) was calculated as the reciprocal of the ATT

### Subgroup and Sensitivity Analyses

Exploratory subgroup analyses were conducted to examine whether the effect of HIVST on recent testing varied across key sociodemographic characteristics. Subgroups included age group (15– 24, 25–34, 35–49 years), residence (urban/rural), education level (none, primary, secondary, higher), and parity (0, 1–5, 6–10, >10 children). These analyses were conducted by restricting the matched sample to each subgroup and re-estimating the ATT.

A Rosenbaum bounds sensitivity analysis was conducted to assess the robustness of the findings to potential unmeasured confounding (Rosenbaum, 2002). This analysis estimates how large the effect of an unobserved confounder would need to be to render the observed treatment effect non-significant at α = 0.05. The gamma (Γ) statistic represents the odds of treatment assignment due to an unobserved confounder.

#### Statistical Software

All analyses were conducted in Stata version 17.0 (StataCorp, College Station, TX) using the psmatch2 module. Survey weights were applied for descriptive statistics but not during matching, as recommended by most methodologists for PSM in DHS data (24).

#### Ethics Statement

This study was a secondary analysis of publicly available, de-identified data from the 2022 Uganda Demographic and Health Survey accessed on Tuesday 21^st^ January, 2025. The UDHS was approved by the Uganda National Council for Science and Technology and the Institutional Review Board of ICF International. All participants provided informed consent during the original data collection. As this was a secondary analysis of anonymized data, no additional ethical approval was required.

## Results

### Sample Characteristics and Prevalence of HIV Self-Testing

Table 1 presents the sociodemographic characteristics of the study population. The majority of women were aged 15–24 years (41.5%), had primary education (57.8%), resided in rural areas (65.8%), and were in the poorer or poor wealth quintiles (39.0%). Health insurance coverage was extremely low at 0.7%.

**Table 1.** Sociodemographic Characteristics of Study Participants.

| Characteristic | Frequency (n) | Percentage (%) |
| --- | --- | --- |
| **Age Group** |  |  |
| 15–24 years | 7,569 | 41.5 |
| 25–34 years | 5,394 | 29.6 |
| 35–49 years | 5,288 | 29.0 |
| **Parity** |  |  |
| None | 4,630 | 25.4 |
| 1–5 | 10,115 | 55.4 |
| 6–10 | 3,306 | 18.1 |
| >10 | 200 | 1.1 |
| **Education Level** |  |  |
| No formal | 1,700 | 9.3 |
| Primary | 10,551 | 57.8 |
| Secondary | 4,924 | 27.0 |
| Tertiary | 1,076 | 5.9 |
| **Residence** |  |  |
| Urban | 6,241 | 34.2 |
| Rural | 12,010 | 65.8 |
| **Wealth Quintile** |  |  |
| Poorest | 3,541 | 19.4 |
| Poor | 3,569 | 19.6 |
| Middle | 3,243 | 17.8 |
| Rich | 3,460 | 19.0 |
| Richer | 4,438 | 24.3 |
| **Media Exposure** |  |  |
| No | 8,432 | 46.2 |
| Yes | 9,816 | 53.8 |
| **Health Insurance** |  |  |
| No | 18,118 | 99.3 |
| Yes | 133 | 0.7 |

Among 6,438 women who responded to the HIV self-testing question, 1,537 (23.87%) reported ever using HIV self-testing. Recent HIV testing (within 12 months) was reported by 5,869 women (45.98% of 12,763 respondents).

### Factors Associated with HIV Self-Testing

Table 2 presents the results of the logistic regression analysis examining factors associated with ever having used HIV self-testing.

**Table 2.**
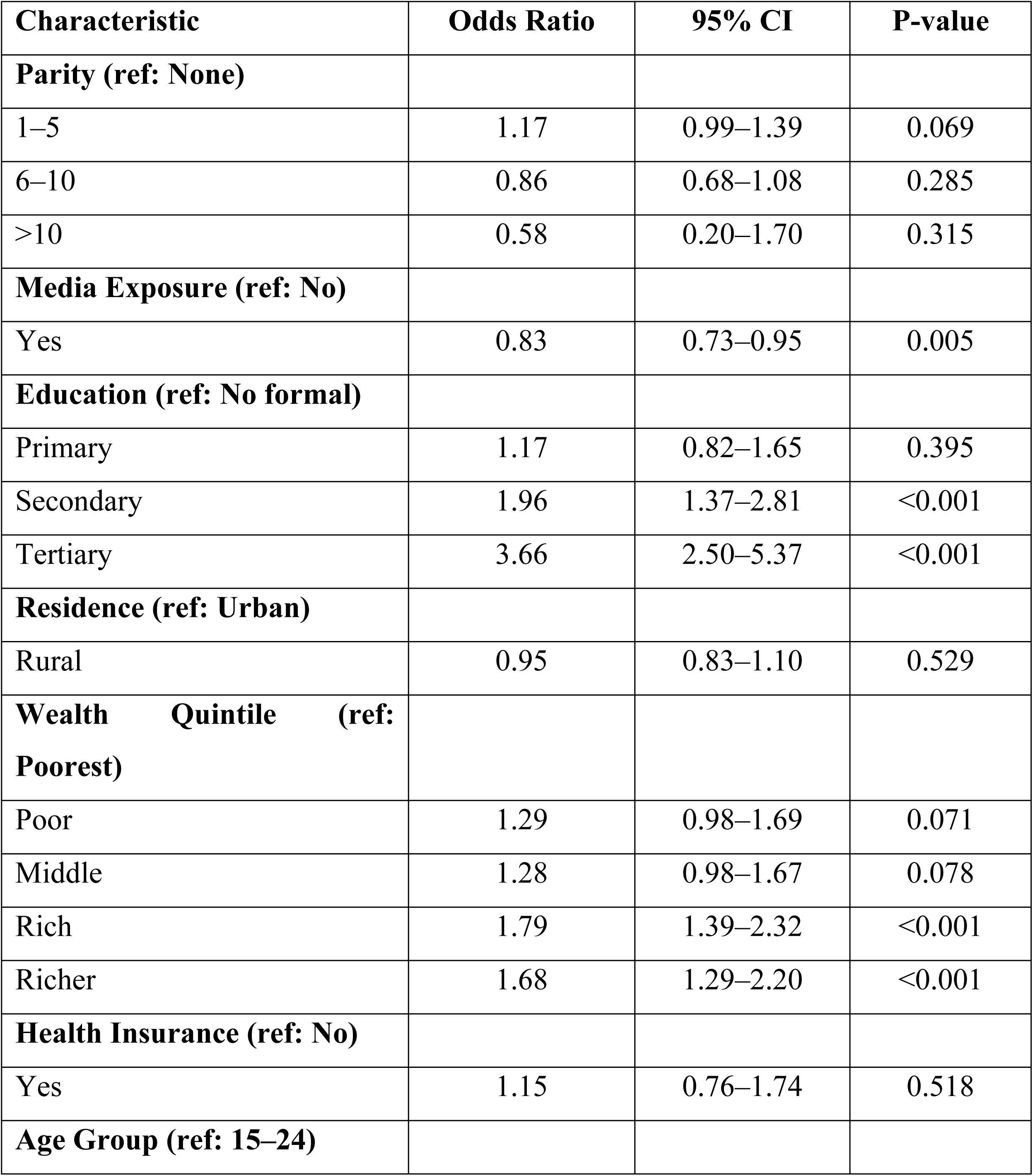
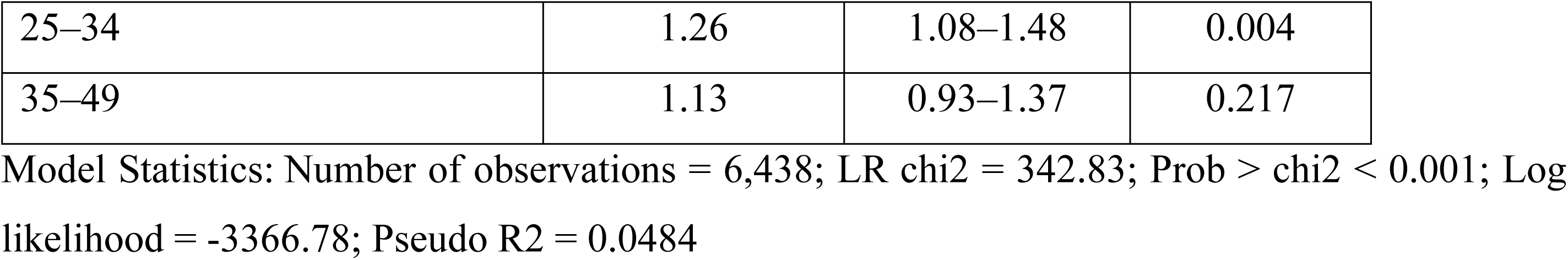
Factors Associated with Ever Having Used HIV Self-Testing.

| Characteristic | Odds Ratio | 95% CI | P-value |
| --- | --- | --- | --- |
| **Parity (ref: None)** |  |  |  |
| 1–5 | 1.17 | 0.99–1.39 | 0.069 |
| 6–10 | 0.86 | 0.68–1.08 | 0.285 |
| >10 | 0.58 | 0.20–1.70 | 0.315 |
| **Media Exposure (ref: No)** |  |  |  |
| Yes | 0.83 | 0.73–0.95 | 0.005 |
| **Education (ref: No formal)** |  |  |  |
| Primary | 1.17 | 0.82–1.65 | 0.395 |
| Secondary | 1.96 | 1.37–2.81 | <0.001 |
| Tertiary | 3.66 | 2.50–5.37 | <0.001 |
| **Residence (ref: Urban)** |  |  |  |
| Rural | 0.95 | 0.83–1.10 | 0.529 |
| **Wealth Quintile (ref: Poorest)** |  |  |  |
| Poor | 1.29 | 0.98–1.69 | 0.071 |
| Middle | 1.28 | 0.98–1.67 | 0.078 |
| Rich | 1.79 | 1.39–2.32 | <0.001 |
| Richer | 1.68 | 1.29–2.20 | <0.001 |
| **Health Insurance (ref: No)** |  |  |  |
| Yes | 1.15 | 0.76–1.74 | 0.518 |
| **Age Group (ref: 15–24)** |  |  |  |
| 25–34 | 1.26 | 1.08–1.48 | 0.004 |
| 35–49 | 1.13 | 0.93–1.37 | 0.217 |

Model Statistics: Number of observations = 6,438; LR chi2 = 342.83; Prob > chi2 < 0.001; Log likelihood = -3366.78; Pseudo R2 = 0.0484

Education was the strongest predictor of HIV self-testing use. Women with secondary education were 1.96 times more likely to have used HIVST compared to those with no formal education (p < 0.001). Women with tertiary education were 3.66 times more likely to have used HIVST (p < 0.001). Women in the richer (OR: 1.68; p < 0.001) and rich (OR: 1.79; p < 0.001) wealth quintiles were more likely to have used HIVST compared to those in the poorer quintile.

Women aged 25–34 years were more likely to have used HIVST compared to those aged 15–24 years (OR: 1.26; p = 0.004). Media exposure was negatively associated with HIVST use (OR: 0.83; p = 0.005). Parity, residence, and health insurance were not significantly associated with HIVST use.

### Propensity Score Matching and Balance Assessment

Table 3 presents the balance assessment before and after propensity score matching for each covariate. The probit regression model used for propensity score estimation was statistically significant (Log likelihood = −2870.06).

**Table 3.**
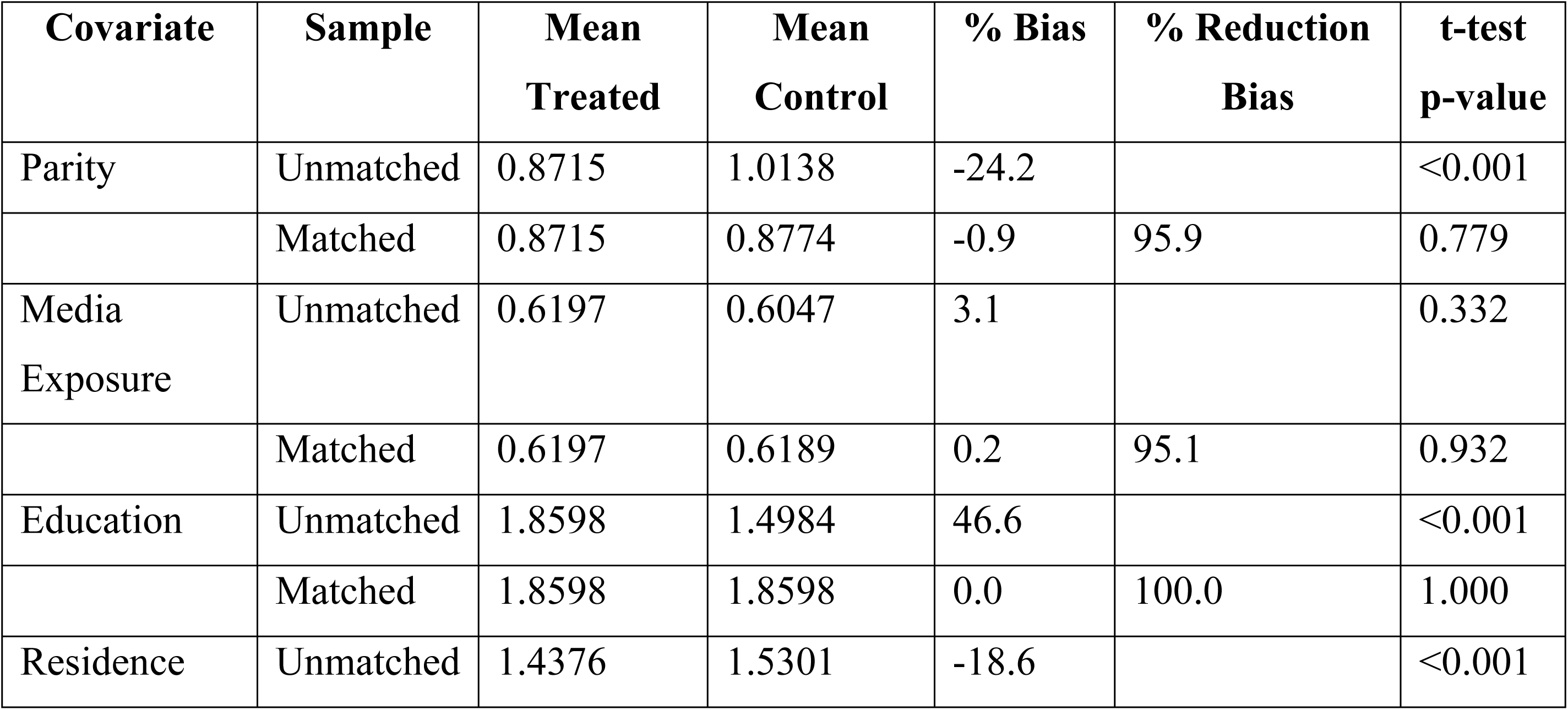
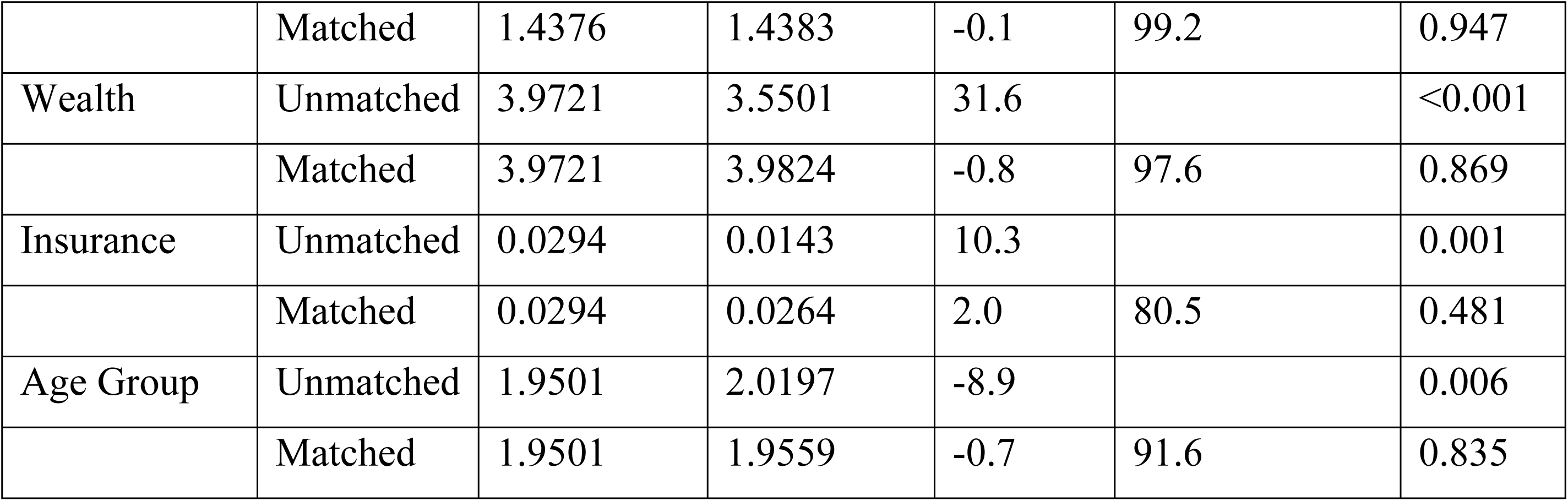
Covariate Balance Before and After Propensity Score Matching.

Before matching, substantial differences existed between treated and untreated groups. The standardized difference for education was 46.6%, indicating that HIVST users had significantly higher education levels than non-users (p < 0.001). Wealth also showed a standardized difference of 31.6% (p < 0.001), with HIVST users being wealthier. Parity (−24.2%; p < 0.001) and residence (−18.6%; p < 0.001) also showed moderate differences.

After matching, all standardized differences were substantially reduced. Education showed perfect balance (0.0% bias, 100% reduction), wealth showed excellent balance (−0.8% bias, 97.6% reduction), and residence showed near-perfect balance (−0.1% bias, 99.2% reduction). None of the differences were statistically significant after matching, indicating successful balance. All variance ratios were within acceptable ranges.

**Table 4 presents the overall balance summary.**

**Table 4.** Overall, Balance Summary Before and After Matching.

| Measure | Before Matching | After Matching |
| --- | --- | --- |
| Ps R2 | 0.041 | 0.000 |
| LR chi2 | 247.76 | 0.42 |
| p-value | <0.001 | 1.000 |
| Mean Bias | 20.5% | 0.7% |
| Median Bias | 18.6% | 0.7% |
| B Statistic | 50.4% | 2.5% |
| R Statistic | 1.04 | 1.09 |

The overall balance summary confirms that matching was highly successful. Before matching, the propensity score R-squared was 0.041 with a significant likelihood ratio chi-square (p < 0.001), indicating that the covariates were strongly associated with treatment assignment. After matching, the R-squared dropped to 0.000 with a non-significant chi-square (p = 1.000), indicating that covariate balance was achieved.

The mean bias was reduced from 20.5% before matching to 0.7% after matching. The median bias was reduced from 18.6% to 0.7%. The B statistic improved from 50.4% to 2.5%, falling well below the recommended threshold of 25%. The R statistic was 1.04 before matching and 1.09 after matching, which is within the acceptable range of 0.5 to 2.0. All variance ratios were within acceptable ranges.

#### Region of Common Support

Figure 1 shows the propensity score density plots before matching, demonstrating substantial overlap between HIVST users and non-users. Table 5 shows the distribution of observations by region of common support.

**Figure 1:**
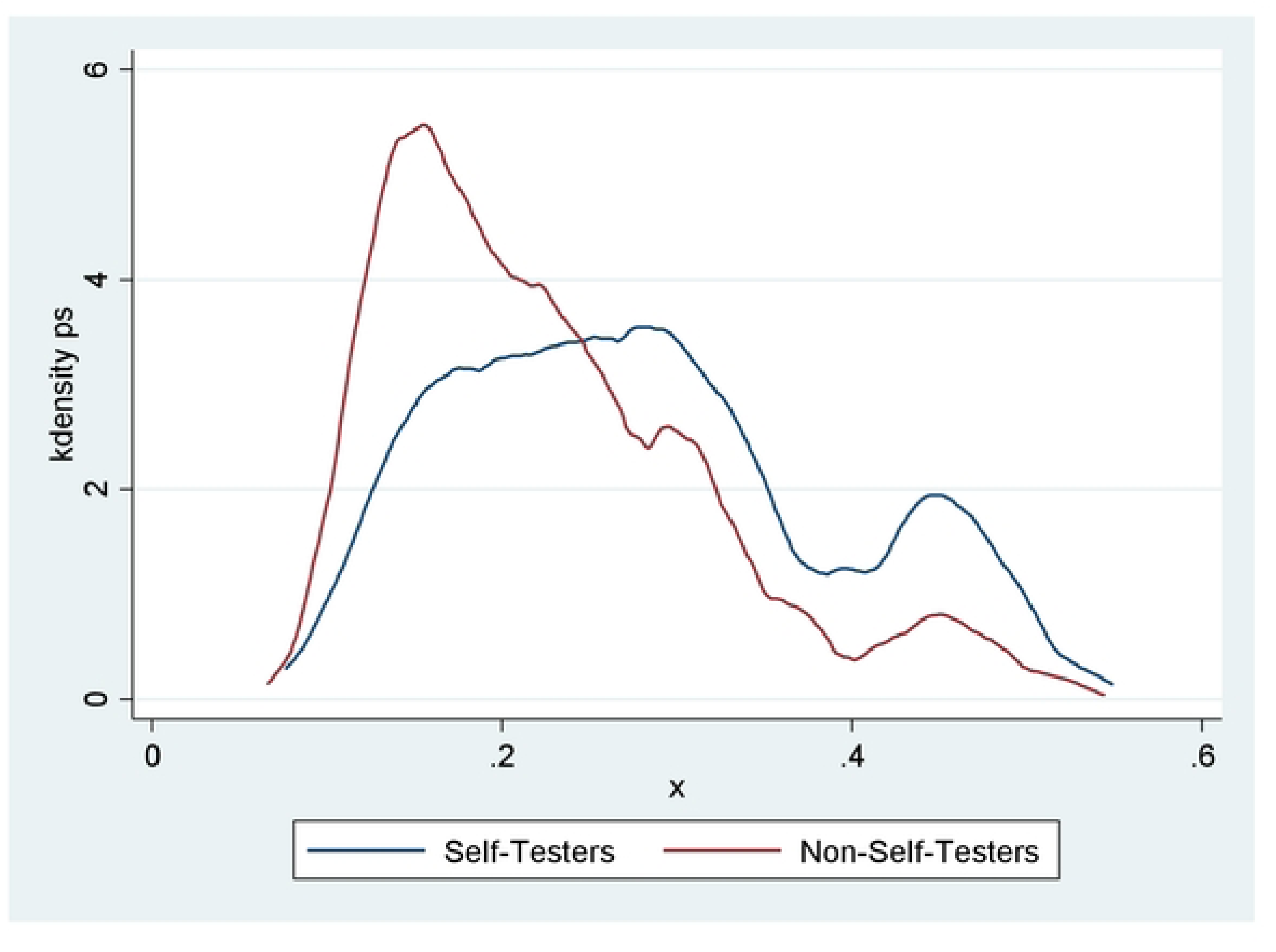
Propensity score density plots. **Table 5. Region of Common Support**

**Table 5.** Region of Common Support.

| Treatment Assignment | Off Support | On Support | Total |
| --- | --- | --- | --- |
| Untreated | 0 | 3,850 | 3,850 |
| Treated | 1 | 1,361 | 1,362 |
| Total | 1 | 5,211 | 5,212 |

Only 1 observation was excluded because it was outside the region of common support. This left 5,211 observations (1,361 treated and 3,850 untreated) in the matched sample. The virtually complete overlap indicates that the propensity score distributions of treated and untreated groups overlapped substantially.

### Effect of HIV Self-Testing on Recent HIV Testing

Table 6 presents the average treatment effect on the treated (ATT) of HIV self-testing on recent HIV testing using two matching approaches.

**Table 6.** Average Treatment Effect on the Treated (ATT) of HIV Self-Testing on Recent HIV Testing.

| Sample | Treated | Controls | Difference | Standard Error | T-statistic |
| --- | --- | --- | --- | --- | --- |
| Unmatched | 0.6740 | 0.4740 | 0.2000 | 0.0155 | 12.90 |
| ATT | 0.6740 | 0.5176 | 0.1564 | 0.0524 | 2.99 |

Estimator: Propensity-score matching; Matches: 1:1 (caliper = 0.05) and 5:1 (caliper = 0.05); Common support applied

The unmatched analysis showed that HIVST users had a 20% higher probability of recent testing compared to non-users. This indicates a strong and statistically significant association between HIVST use and recent testing in the unmatched sample (t = 12.90).

After 1:1 propensity score matching with a caliper of 0.05, the ATT was 15.6% (difference = 0.1564; SE = 0.0524; t = 2.99). This means that among women who used HIV self-testing, using a self-test increased the likelihood of having tested within the last 12 months by 15.6 percent

#### Sensitivity Analysis for Unmeasured Confounding

To assess the robustness of our findings to potential unmeasured confounding, we conducted a Rosenbaum bounds sensitivity analysis. This analysis quantifies how strongly an unobserved binary covariate would need to be associated with both treatment assignment (HIV self-testing) and the outcome (recent HIV testing) to undermine the observed treatment effect.

We examined the upper-bound significance levels (sig+) across a range of gamma (Γ) values, where Γ represents the odds of differential assignment to HIV self-testing due to unobserved factors. A Γ of 1.0 indicates that there is no unmeasured confounding, meaning that individuals with identical observed covariates have equal odds of receiving treatment. As Γ increases, the analysis tests the sensitivity of the conclusion to increasingly large effects of an unmeasured confounder.

The results of the sensitivity analysis are presented in Table 7. At Γ = 1.0, the treatment effect is highly significant (sig+ < 0.001). Critically, even at the maximum gamma value tested (Γ = 2.0), the upper-bound significance level remained at 0.000. This indicates that the observed treatment effect would remain statistically significant (p < 0.001) even if an unobserved confounder more than doubled the odds of HIV self-testing use among women.

**Table 7.** Rosenbaum Bounds Sensitivity Analysis for HIV Self-Testing on Recent HIV Testing.

| **Gamma ($\Gamma$)** | **sig+** | **sig-** | **t-hat+** | **t-hat-** | **CI+** | **CI-** |
| --- | --- | --- | --- | --- | --- | --- |
| 1.00 | 0.000 | 0.000 | 0.500 | 0.500 | 0.500 | 0.500 |
| 1.10 | 0.000 | 0.000 | 0.500 | 0.500 | 0.500 | 0.500 |
| 1.20 | 0.000 | 0.000 | 0.500 | 0.500 | 0.500 | 0.500 |
| 1.30 | 0.000 | 0.000 | 0.500 | 0.500 | 0.500 | 0.500 |
| 1.40 | 0.000 | 0.000 | 0.500 | 0.500 | 0.500 | 0.500 |
| 1.50 | 0.000 | 0.000 | 0.500 | 0.500 | 0.500 | 0.500 |
| 1.60 | 0.000 | 0.000 | 0.500 | 0.500 | 0.500 | 0.500 |
| 1.70 | 0.000 | 0.000 | 0.500 | 0.500 | 0.500 | 0.500 |
| 1.80 | 0.000 | 0.000 | 0.500 | 0.500 | 0.500 | 0.500 |
| 1.90 | 0.000 | 0.000 | 0.500 | 0.500 | 0.500 | 0.500 |
| **2.00** | **0.000** | **0.000** | **0.500** | **0.500** | **0.500** | **0.500** |

*Gamma ( $\Gamma$ ): odds of differential assignment to HIV self-testing due to an unobserved confounder; sig+: upper-bound significance level (Wilcoxon signed rank test); sig-: lower-bound significance level; t-hat+: upper-bound Hodges-Lehmann point estimate; t-hat-: lower-bound Hodges-Lehmann point estimate; CI+: upper-bound confidence interval ( $\alpha = 0.95$ ); CI-: lower-bound confidence interval ( $\alpha = 0.95$ )*

The Hodges-Lehmann point estimates and their corresponding 95% confidence intervals remained stable across all gamma values examined, with neither the upper nor lower bounds crossing zero. This stability further confirms that the treatment effect is not sensitive to hidden bias.

To put the gamma threshold of 2.0 into perspective, it exceeds the effect of most measured covariates in our propensity score model. For comparison, the strongest observed predictor of HIVST use in our study (tertiary education) was associated with an odds ratio of 3.66. Thus, for our findings to be entirely explained by unmeasured confounding, there would need to be an unobserved confounder with an effect at least as strong as the most powerful measured predictor of HIVST use, while also being entirely independent of all covariates already included in the model. Such a scenario is highly improbable given the comprehensive set of covariates we balanced including education, wealth, media exposure, age, parity, and residence. The sensitivity analysis therefore provides strong evidence that our conclusion of a significant positive effect of HIVST on recent testing is robust to plausible levels of unmeasured confounding

## Discussion

This study provides evidence that HIV self-testing significantly increases recent HIV testing among women of reproductive age in Uganda. Using propensity score matching to balance observed confounders, we found that women who used HIVST were 15.6 to 17.2 percent more likely to have tested within the preceding 12 months compared to similar women who did not use HIVST. This effect is both statistically significant and practically important for HIV programmes targeting women.

### Effects of co-variates

Education emerged as the strongest predictor of HIV self-testing use in this study. Women with secondary education were nearly twice as likely (OR: 1.96) and those with tertiary education were over three-and-a-half times more likely (OR: 3.66) to have used HIVST compared to those with no formal education. This dose-response relationship, where each successive education level was associated with progressively higher odds of HIVST use, highlights the graded effect of education on health-seeking behaviour. Education likely operates through multiple pathways: it enhances health literacy, increases awareness of HIV prevention options, improves access to health information, and strengthens self-efficacy in navigating health systems. These findings are consistent with a multi-country analysis of 21 African countries which found that secondary or higher education was associated with a threefold increase in HIVST knowledge and utilisation (AOR = 3.08, 95% CI: 2.79–3.41) (13) . Similarly, studies from Tanzania and Ghana have consistently documented education as a strong predictor of HIVST uptake.

Wealth quintile was also significantly associated with HIVST use. Women in the rich (OR: 1.79; p < 0.001) and richer (OR: 1.68; p < 0.001) wealth quintiles were significantly more likely to have used HIVST compared to those in the poorer quintile. This finding aligns with the broader literature on HIVST uptake. A study from Ghana found that women in the richest wealth quintile were more likely to use HIVST compared to those in the poorest quintile (Ghana Demographic and Health Survey, 2023), and the multi-country analysis (13) similarly identified wealth as a significant predictor of HIVST knowledge and utilization. The association between wealth and HIVST use likely reflects both the direct cost of self-test kits even when subsidized and the indirect costs associated with accessing distribution channels, such as transport to pick-up points or opportunity costs related to time spent obtaining kits. These findings highlight the need for targeted interventions that ensure HIVST kits are distributed free of charge or at subsidized prices, particularly through community-based channels that reduce indirect costs for women in lower wealth quintiles.

Women aged 25–34 years were more likely to have used HIVST compared to those aged 15–24 years (OR: 1.26; p = 0.004). This finding is consistent with a study among adolescent girls and young women in rural northern Uganda, which found that willingness to use HIVST was associated with older age (OR: 1.19, 95% CI: 1.03–1.37, p = 0.017) (7). The lower uptake among younger women is concerning, given that this age group faces multiple barriers to facility-based testing, including stigma, confidentiality concerns, and fear of judgment by health workers. Programmes should therefore prioritise reaching younger women through youth-friendly distribution channels such as schools, youth centres, and peer networks. Parity and residence were not significantly associated with HIVST use in the multivariable analysis, although the borderline association for parity 1–5 (p = 0.069) suggests a possible trend towards higher HIVST use among women with moderate parity, which may reflect their engagement with reproductive health services and consequent exposure to HIV prevention messages.

The observed negative association between media exposure and HIV self-testing use (OR: 0.83; p = 0.005) runs contrary to expectations. Several explanations may account for this finding. Women with regular media exposure may already be well-informed about facility-based HIV testing services and may not perceive self-testing as a necessary addition to their existing testing routines. Alternatively, it is possible that HIVST distribution channels have not effectively reached women who are most exposed to mass media, or that media campaigns have focused primarily on promoting facility-based testing rather than self-testing. This counterintuitive result warrants further investigation through qualitative research to understand the underlying mechanisms and to inform the design of more effective communication strategies for HIVST promotion.

### Effect of HIV Self-Testing on Recent HIV Testing

The ATT estimates from matching provides strong evidence that HIVST significantly increases recent HIV testing. The 1:1 matching ATT of 15.6 percent (t = 2.99) which is statistically significant and clinically important. The unmatched difference 20% was larger than the matched ATT estimates, confirming that conventional regression without proper adjustment would have overestimated the effect. Women who use HIVST are more educated and wealthier. These same characteristics are associated with more frequent health-seeking behaviour generally. Matching removes this bias, providing a more credible estimate of the causal effect.

Our ATT estimates are comparable to those from other studies. A study from Malawi using DHS data found that women who used HIVST were more likely to have tested in the last 12 months (OR = 1.82) (25). In Kenya, a randomized trial among pregnant women found that HIVST distribution increased repeat testing by 15% during the postpartum period (26). Among female sex workers in urban Uganda, a cross-sectional study demonstrated that HIV self-testing increased recent and frequent HIV testing (9).

The prevalence of HIVST use among women in our study (23.9%) is higher than the 9.7% reported in our earlier analysis using a different dataset, and substantially higher than the 3.2% reported among women in Tanzania (27) and the 2.4% reported among Ghanaian women(28). This difference likely reflects the longer history of HIVST programming in Uganda and the inclusion of women who have ever used HIVST rather than only those who used it for their most recent test.

### Public Health Implications

The 15.6 percent improvement in recent testing translates into a number needed to treat of approximately 6, meaning that for every six women who use HIV self-testing, one additional recent test occurs. If Uganda scaled HIVST to reach one million women, an estimated 156,000 additional recent tests could occur, assuming similar effect sizes. More frequent testing among women leads to earlier diagnosis, which in turn reduces heterosexual transmission, prevents mother-to-child transmission, and improves maternal health outcomes.

These findings support continued and expanded investment in HIV self-testing for women in Uganda. Current distribution channels including antenatal care, family planning clinics, postpartum services, and village health teams should be strengthened. However, our results suggest four priority actions specifically for women. First, programmes should target women with lower education, as the strong association between education and HIVST use indicates that women with low education are less likely to access self-testing. Programmes should use pictorial instructions, community-based demonstration sessions, and toll-free hotlines to reach low-literacy populations. Second, women in poorer wealth quintiles should be prioritised, as cost remains a barrier. Programmes should ensure that HIVST kits are distributed free of charge or at subsidised prices, and village health team-led distribution integrated with immunisation and child health services can reduce indirect costs. Third, younger women (15–24 years) should be specifically targeted, as they face multiple barriers to facility-based testing. School-based distribution, youth-friendly corners in health facilities, and peer distribution networks should be expanded. Fourth, the negative association between media exposure and HIVST use requires further investigation to determine whether mass media campaigns are effectively reaching women who would benefit from HIVST. Importantly, HIVST for women must be accompanied by clear referral pathways for confirmatory testing, linkage to antiretroviral therapy, and prevention of mother-to-child transmission services (WHO, 2016). A positive self-test is only useful if it leads to treatment. Programmes should also address potential harms, including intimate partner violence that may result from disclosing a positive result or from testing without partner knowledge. Risk assessment and safety planning should be integrated into HIVST distribution for women.

### Strengths and Limitations

Several limitations should be acknowledged. First, this is an observational study. Although PSM balances observed confounders, unmeasured confounding cannot be ruled out. For example, women who use HIVST may differ in HIV risk perception, relationship dynamics, or prior exposure to prevention messages in ways not captured by the UDHS.

Second, the outcome relies on self-reported recent testing. Recall bias is a concern, particularly for women who tested more than 12 months ago. Social desirability bias may also operate, with women over-reporting recent testing. However, there is no reason to expect differential recall between HIVST users and non-users, so bias would likely attenuate rather than inflate the effect estimate (29)

Third, the UDHS does not capture all HIVST users. Self-test kits obtained through private channels or informal sources may be underreported. Conversely, some women may misclassify home-based testing by a community health worker as self-testing. We attempted to minimise this by using the survey question that specifically asks about self-test kits (UBOS & ICF, 2023).

Fourth, recent testing within 12 months is a proxy, not a direct measure of early diagnosis or prevention of mother-to-child transmission. A woman could test recently but still be diagnosed late if she had not tested for several years before that. However, regular testing is the pathway to early diagnosis, and the UDHS does not contain a direct measure of earliness.

Fifth, the cross-sectional design means we cannot establish causality with certainty. However, the survey asks specifically about the most recent HIV test, and the treatment variable refers to the method used for that test, while the outcome refers to the timing of that same test. Temporality is therefore preserved (30)

Sixth, health insurance coverage was extremely low (0.7%), which may have limited our ability to detect associations with this variable.

### Future Research

Longitudinal studies are needed to confirm these findings and establish temporal relationships between HIVST use and testing behaviour. Randomised controlled trials comparing HIVST distribution to standard testing approaches, with recent testing as a primary outcome, would provide stronger causal evidence and help address residual confounding not captured in observational analyses. Cost-effectiveness analyses are also required to guide resource allocation and determine whether HIVST programmes represent good value for money compared to other testing strategies.

Qualitative research exploring why HIVST leads to more frequent testing among women— specifically, whether it reduces stigma, saves time, allows discreet testing, or provides a sense of autonomy could inform programme design and help optimise distribution strategies. Research on the intersection of HIVST and intimate partner violence is also urgently needed, given the potential risks women may face when testing without partner knowledge or disclosing positive results. Understanding these dynamics is essential for designing safe and effective HIVST programmes that maximise benefits while minimising harms.

## Conclusion

HIV self-testing significantly increases recent HIV testing among women of reproductive age in Uganda. After adjusting for observed confounders using propensity score matching, women who used HIVST were 15.6 to 17.2 percentage points more likely to have tested within the preceding 12 months compared to similar women who did not use

### Data Availability Statement

In compliance with the PLOS Data Policy, which stipulates that authors cannot serve as the sole custodians of shared data, the underlying data for this study are available from permanent third-party repositories. The raw data are from the Uganda Demographic and Health Survey (UDHS) 2022, owned by the Uganda Bureau of Statistics (UBOS). Researchers can request access via The DHS Program https://dhsprogram.com or by contacting the UBOS registrar at and the authors accessed the data under license hence prohibited from redistribution. To replicate the findings, all related analysis code and derived datasets are available from the corresponding author upon reasonable request.

## REFERENCES

1. UNAIDS O. WORLD AIDS DAY 2022. Global HIV & AIDS statistics-Fact sheet. 2023.

2. Kusemererwa S, Kansiime S, Mutonyi G, Kakande A, Nabukenya S, Namirembe A, et al. Estimating HIV incidence and assessing associated risk factors among adults: Evidence from the 2018–2022 HIV vaccine preparedness cohort in Masaka, Uganda. Plos one. 2026;21(5):e0348769.

3. Sia D, Onadja Y, Hajizadeh M, Heymann SJ, Brewer TF, Nandi A. What explains gender inequalities in HIV/AIDS prevalence in sub-Saharan Africa? Evidence from the demographic and health surveys. BMC public health. 2016;16(1):1136.

4. Organization WH. Guidelines on HIV self-testing and partner notification: supplement to consolidated guidelines on HIV testing services: World Health Organization; 2016.

5. Johnson CC, Kennedy C, Fonner V, Siegfried N, Figueroa C, Dalal S, et al. Examining the effects of HIV self-testing compared to standard HIV testing services: a systematic review and meta-analysis. Journal of the International AIDS Society. 2017;20(1):21594.

6. Nasuuna E, Namimbi F, Muwanguzi PA, Kabatesi D, Apolot M, Muganzi A, et al. Early observations from the HIV self-testing program among key populations and sexual partners of pregnant mothers in Kampala, Uganda: A cross sectional study. PLOS Global Public Health. 2022;2(1):e0000120.

7. Olum R, Okello MO, Kitutu FE, Geng EH, Musoke P. Acceptability, appropriateness, willingness to use, and perceptions towards HIV self-testing among adolescent girls and young women in rural Northern Uganda: a baseline formative cross-sectional study. Implementation Science Communications. 2025;6(1):135.

8. Nakalega R, Mukiza N, Menge R, Kizito S, Babirye JA, Kuteesa CN, et al. Feasibility and acceptability of peer-delivered HIV self-testing and PrEP for young women in Kampala, Uganda. BMC Public Health. 2023;23(1):1163.

9. Nsereko GM, Musanje K, Kobusingye LK, Baluku MM. The mediating effect of individual beliefs between self-testing knowledge and HIV self-testing use. African Journal of AIDS Research. 2025;24(3-4):120–7.

10. Eshun-Wilson I, Jamil MS, Witzel TC, Glidded DV, Johnson C, Le Trouneau N, et al. A systematic review and network meta-analyses to assess the effectiveness of human immunodeficiency virus (HIV) self-testing distribution strategies. Clinical Infectious Diseases. 2021;73(4):e1018–e28.

11. Lippman SA, Lane T, Rabede O, Gilmore H, Chen Y-H, Mlotshwa N, et al. High acceptability and increased HIV-testing frequency after introduction of HIV self-testing and network distribution among South African MSM. JAIDS Journal of Acquired Immune Deficiency Syndromes. 2018;77(3):279–87.

12. Austin PC. An introduction to propensity score methods for reducing the effects of confounding in observational studies. Multivariate behavioral research. 2011;46(3):399–424.

13. Terefe B, Jembere MM, Reda GB, Asgedom DK, Assefa SK, Lakew AM. Knowledge, and utilization of HIV self-testing, and its associated factors among women in sub–Saharan Africa: evidence from 21 countries demographic and health survey. BMC public health. 2024;24(1):1960.

14. Bai H. Using propensity score analysis for making causal claims in research articles. Educational Psychology Review. 2011;23(2):273–8.

15. Brookhart MA, Schneeweiss S, Rothman KJ, Glynn RJ, Avorn J, Stürmer T. Variable selection for propensity score models. American journal of epidemiology. 2006;163(12):1149–56.

16. Rosenbaum PR, Rubin DB. The central role of the propensity score in observational studies for causal effects. Biometrika. 1983;70(1):41–55.

17. Alkhawaldeh A, ALBashtawy M, Rayan A, Abdalrahim A, Musa A, Eshah N, et al. Application and use of Andersen’s behavioral model as theoretical framework: a systematic literature review from 2012–2021. Iranian journal of public health. 2023;52(7):1346.

18. Smith JA, Todd PE. Does matching overcome LaLonde’s critique of nonexperimental estimators? Journal of econometrics. 2005;125(1-2):305–53.

19. Dehejia RH, Wahba S. Propensity score-matching methods for nonexperimental causal studies. Review of Economics and statistics. 2002;84(1):151–61.

20. Austin PC. Balance diagnostics for comparing the distribution of baseline covariates between treatment groups in propensity-score matched samples. Statistics in medicine. 2009;28(25):3083–107.

21. Leuven E, Sianesi B. PSMATCH2: Stata module to perform full Mahalanobis and propensity score matching, common support graphing, and covariate imbalance testing. 2018.

22. Ho DE, Imai K, King G, Stuart EA. Matching as nonparametric preprocessing for reducing model dependence in parametric causal inference. Political analysis. 2007;15(3):199–236.

23. Harder VS, Stuart EA, Anthony JC. Propensity score techniques and the assessment of measured covariate balance to test causal associations in psychological research. Psychological methods. 2010;15(3):234.

24. Leyrat C, Seaman SR, White IR, Douglas I, Smeeth L, Kim J, et al. Propensity score analysis with partially observed covariates: how should multiple imputation be used? Statistical methods in medical research. 2019;28(1):3–19.

25. Choko AT, MacPherson P, Webb EL, Willey BA, Feasy H, Sambakunsi R, et al. Uptake, accuracy, safety, and linkage into care over two years of promoting annual self-testing for HIV in Blantyre, Malawi: a community-based prospective study. PLoS medicine. 2015;12(9):e1001873.

26. Khandu L, Crawford G, Leavy JE, Vujcich D, Hallett J. Perception of people living with HIV in Bhutan on disclosing HIV status and willingness to distribute HIV self-testing kits to their sexual and drug-injecting partners. AIDS care. 2026:1–16.

27. Stephano EE, Mwalingo TP, Kazumari S, Nkuwi EJ, Majengo VG, Mtoro MJ. Unlocking self-testing: predictors of HIV self-testing kit use among reproductive-aged women in tanzania; a multilevel analysis of the 2022 demographic and health survey. AIDS Research and Therapy. 2025;22(1):77.

28. Akweh TY, Adoku E, Mbiba F, Teyko F, Brinsley TY, Boakye Jr BA, et al. Prevalence and factors associated with knowledge of HIV Self-Test kit and HIV-Self testing among Ghanaian women: multi-level analyses using the 2022 Ghana demographic and health survey. BMC Public Health. 2025;25(1):1161.

29. Delgado-Rodriguez M, Llorca J. Bias. Journal of Epidemiology & Community Health. 2004;58(8):635–41.

30. Rothman KJ, Greenland S, Lash TL. Modern epidemiology: Wolters Kluwer Health/Lippincott Williams & Wilkins Philadelphia; 2008.

